# Can increasing years of schooling reduce type 2 diabetes (T2D)?—Evidence from a Mendelian randomization of T2D and 10 of its risk factors

**DOI:** 10.1101/2020.02.05.20020701

**Authors:** Charleen D. Adams, Brian B. Boutwell

## Abstract

A focus in recent decades has involved examining the potential causal impact of educational attainment (schooling years) on a variety of disease and life-expectancy outcomes. Numerous studies have broadly revealed a link suggesting that as years of formal schooling increase so too does health and wellbeing; however, it is unclear whether the associations are causal. Here we use Mendelian randomization, an instrumental variables technique, to probe whether more years of schooling are causally linked to type 2 diabetes (T2D) and 10 of its risk factors. The results reveal a protective effect of more schooling years against T2D (odds ratio=0.39; 95% confidence interval: 0.26, 0.58; *P*=3.89 × 10^−06^), which might be mediated in part by more years of schooling being protective against the following: having a first-degree relative with diabetes, being overweight, and having high blood pressure, higher levels of circulating triglycerides, and lower levels of HDL cholesterol. More schooling years had no effect on risk for gestational diabetes or polycystic ovarian syndrome and was associated with a decreased likelihood of moderate physical activity. These findings imply that strategies to retain adults in higher education may help reduce the risk for a major source of metabolic morbidity and mortality.

---

Tacit to most epidemiological research is a desire to infer whether an environmental exposure impacts some outcome in a causal fashion. A particular area of focus in recent decades has involved examining the impact of educational attainment (years of schooling) on a variety of disease and life expectancy outcomes^1^. Numerous studies have broadly revealed a strong statistical association suggesting that as the years of formal schooling increase so too does health and wellbeing ^2^. Indeed, educational attainment has been associated with diverse mental and physical health outcomes, including depression, cancer incidence, heart disease, and diabetes^1^.

Entangled in this line of inquiry (and all of social science research), however, is a concern about the evidence for causal inference open to scholars^3^. With regard to the associations between educational attainment and health outcomes, Montez and Friedman (2015) caution: “Studies such as those highlighted above often implicitly assume that educational attainment has a causal influence on adult health; however, this assumption has long been challenged. If the assumption is incorrect then investing in education policies and schooling systems may waste government spending and not manifest in improved population health” (p.1)^1^. To be sure, there is emergent evidence utilizing quasi-experimental and natural-experimental designs which suggest some causal effects may exist in some contexts for educational attainment on health outcomes^2^. Yet, there remains an overall dearth of evidence utilizing designs admitting of stronger causal inference capabilities.

More recently, scholars have begun utilizing data gleaned from large genomic consortia and publicly available genome wide association (GWA) studies to construct instrumental variables comprised of trait-relevant single-nucleotide polymorphisms (SNPs). When certain assumptions (discussed below) are satisfied in the data, it is possible to investigate whether some type of modifiable risk or protective factor causally impacts some outcome^4^. Known as Mendelian Randomization (MR), this variant of instrumental variable analysis has been increasingly widely applied to a variety of medical and epidemiological outcomes^5^. In the current study, we apply MR modeling strategies to zoom in on whether educational attainment plays a causal role in the prevention of one of society’s most pressing public-health challenges: type 2 diabetes (T2D) and 10 of its risk factors.

## Results

### T2D

A strong protective effect against T2D is observed for more Education Years (odds ratio, OR, for T2D per SD increase in Education Years): IVW estimate 0.39; 95% confidence interval (CI): 0.26, 0.58; *P*=3.89 × 10^−06^). The sensitivity estimators aligned in direction and magnitude of effects with the IVW’s estimate, and the MR-Egger intercept test indicated no evidence for directional pleiotropy. (Since this is also the case for all the tests – none showed evidence for direction pleiotropy with the MR-Egger intercept test, this statement will not be repeated for the remaining results).

### Sibling, mother, and father with diabetes

Small protective effects against having a sibling, mother, or father with diabetes are observed for more Education Years (ORs for a first-degree relative with diabetes per SD increase in Education Years): sibling IVW estimate 0.97; 95% CI: 0.96, 0.98; *P*=4.23 × 10^−11^); mother IVW estimate 0.97; 95% CI: 0.96, 0.98; *P*=6.66 × 10^−7^); father IVW estimate 0.98; 95% CI: 0.97, 0.99; *P*=0.0008. The sensitivity estimators aligned in direction and magnitude of effects with the IVW’s estimate.

### Overweight status

A strong protective effect against being overweight is observed for more Education Years (OR for being overweight per SD increase in Education Years): IVW estimate 0.60; 95% CI: 0.51, 0.72; *P*=1.01 × 10^−08^). The sensitivity estimators mostly aligned in direction and magnitude of effects with the IVW’s estimate, with a slightly more protective effect observed for the weighted mode estimator.

### Physical activity

A strong protective effect against performing the most amount of moderate physical activity is observed for more Education Years (OR for the highest level of moderate physical activity compared to all other amounts of moderate physical activity per SD increase in Education Years): IVW estimate 0.77; 95% CI: 0.71, 0.84; *P*=1.08 × 10^−08^). The sensitivity estimators varied in the magnitude of their effects, which might indicate unwanted pleiotropy.

### High blood pressure

A modest protective effect against having high blood pressure is observed for more Education Years (OR for high blood pressure per SD increase in Education Years): IVW estimate 0.94; 95% CI: 0.92, 0.96; *P*=2.49 × 10^−10^). The sensitivity estimators aligned in direction and magnitude of effects with the IVW’s estimate.

### Gestational diabetes and polycystic ovarian syndrome

There were null effects for the influence of more Education Years on gestational diabetes and polycystic ovarian syndrome (OR for each per SD increase in Education Years): IVW estimate 1.00; 95% CI: 1.00, 1.00; gestational diabetes, *P*=0.1705; polycystic ovarian syndrome, *P*=0.2844. The sensitivity estimators aligned in direction and magnitude of effects with the IVW’s estimate: all = 1.

### HDL levels

An increase in HDL levels were observed for more Education Years (beta estimate per SD increase in Education Years): IVW estimate 0.14; 95% CI: 0.06, 0.22; *P*=0.0009). The sensitivity estimators varied in the magnitude of effects, indicating the potential for some unwanted pleiotropy.

### Triglyceride levels

A decrease in triglyceride levels were observed for more Education Years (beta estimate per SD increase in Education Years): IVW estimate −0.19; 95% CI: −0.27, −0.11; *P*=3.34 × 10^−06^). The sensitivity estimators aligned in direction and magnitude of effects with the IVW’s estimate.

## Discussion

We observed a protective effect of Education Years against T2D, which might be mediated in part by more years of schooling being protective against the following: having a first-degree relative with diabetes, being overweight, and having high blood pressure, higher levels of circulating triglycerides, and lower levels of HDL cholesterol. These findings comport with another MR study that examined education and diabetes with UK Biobank data. Davies *et al*. (2018) observed that leaving secondary school at an older age was causally protective against diabetes^7^. Their study differed from the present one in that ours examined education inclusive of college—Davies *et al*. (2018) focused on education up to college. Here, we document that the protective effect of education extends beyond schooling in adolescence. Years of schooling after high school decrease the chance of T2D.

In the present study, more years of schooling had no effect on risk for gestational diabetes or polycystic ovarian syndrome and was associated with a decreased likelihood of moderate physical activity. Regarding the later, another recent MR study found little evidence that more education increased vigorous physical activity^8^. Thus, it seems unlikely that the protective effect of Education Years against T2D occurs through an influence on physical activity.

The protective effect against having a first-degree relative with diabetes is intriguing. Several recent studies have documented that there is a bidirectional causal relationship between fluid intelligence and years of schooling^9,10^. While having higher fluid intelligence may causally impact more years of schooling, the magnitude of the effect for more years of schooling increasing fluid intelligence is comparatively larger: that is, the impact of Education Years on intelligence is more than two-fold greater than the impact of intelligence on Education Years^9,10^. Like educational attainment, which is sometimes treated as a proxy for cognitive ability, being brighter is protective against an array of negative health-outcomes^11^. This means that it is possible that intelligence is confounding the present findings, especially those pertaining to a protective effect of more years of schooling against having a first-degree relative with diabetes. However, due to the durable influence of educational attainment on intelligence, it is also conceivable that those with more education positively influence their family members in ways that reduce risk for T2D.

One limitation for the analyses of Education Years on a first-degree relative with diabetes is that it is possible that some cases of type 1 diabetes were included, since the UK Biobank questions that captured the measure for illnesses of relatives asked about “diabetes” – not specifically about T2D. However, the influence for this is expected to be minimal, since more than 90% of adults with diabetes have T2D^12^.

The primary limitation of the present study is one that all MR studies are liable to: unwanted horizontal pleiotropy. However, the most logical pleiotropic confounder—intelligence—is one that is influenced by Education Years. Moreover, most of the sensitivity screens for possible violations to the MR assumptions revealed little evidence for distortions due to pleiotropy. The exceptions are for HDL levels and physical activity, for which there was enough variability across the sensitivity estimators to view their results with more caution. A strength of our study worth mentioning is that it leveraged the power of 11 large GWA studies to examine these complexly woven traits.

The public-health relevance of the bidirectional causal relationship between intelligence and Education Years cannot be overstated, however. If the present findings primarily reflect the benefits of higher cognitive ability—which they could—then whether Education Years influences cognitive ability determines interventional strategies. Because Education Years increases cognitive ability, public-health efforts to retain people in higher education may be warranted as part of a developing arsenal to help limit and even prevent the staggeringly deleterious effects of T2D. The message is the same, importantly, even if intelligence is not the driving force in the current study. Whatever it is about the landscape of higher education, more years of schooling appears to help reduce the risk for a major source of metabolic morbidity and mortality.

## Methods

### Conceptual approach

MR is an analytic, instrumental variables technique that capitalizes on Mendel’s Laws of Inheritance, genotype assignment at conception, and pleiotropy (genes influencing more than one trait) for causal inference^13–15^.

MR uses genetic variants strongly associated with traits of interest as opposed to the observed traits themselves in models. By relying on the random assortment of alleles (Mendel’s Laws) and the temporal assignment of genotype at conception, MR avoids most sources of confounding and reverse causation that distort causal estimates in observational studies. In two-sample MR, summary statistics are pulled from two genome-wide association (GWA) studies. These summary statistics are the data sources for two-sample MR^4,6,16–19^ (**Figure 1**).

**Figure 1.**
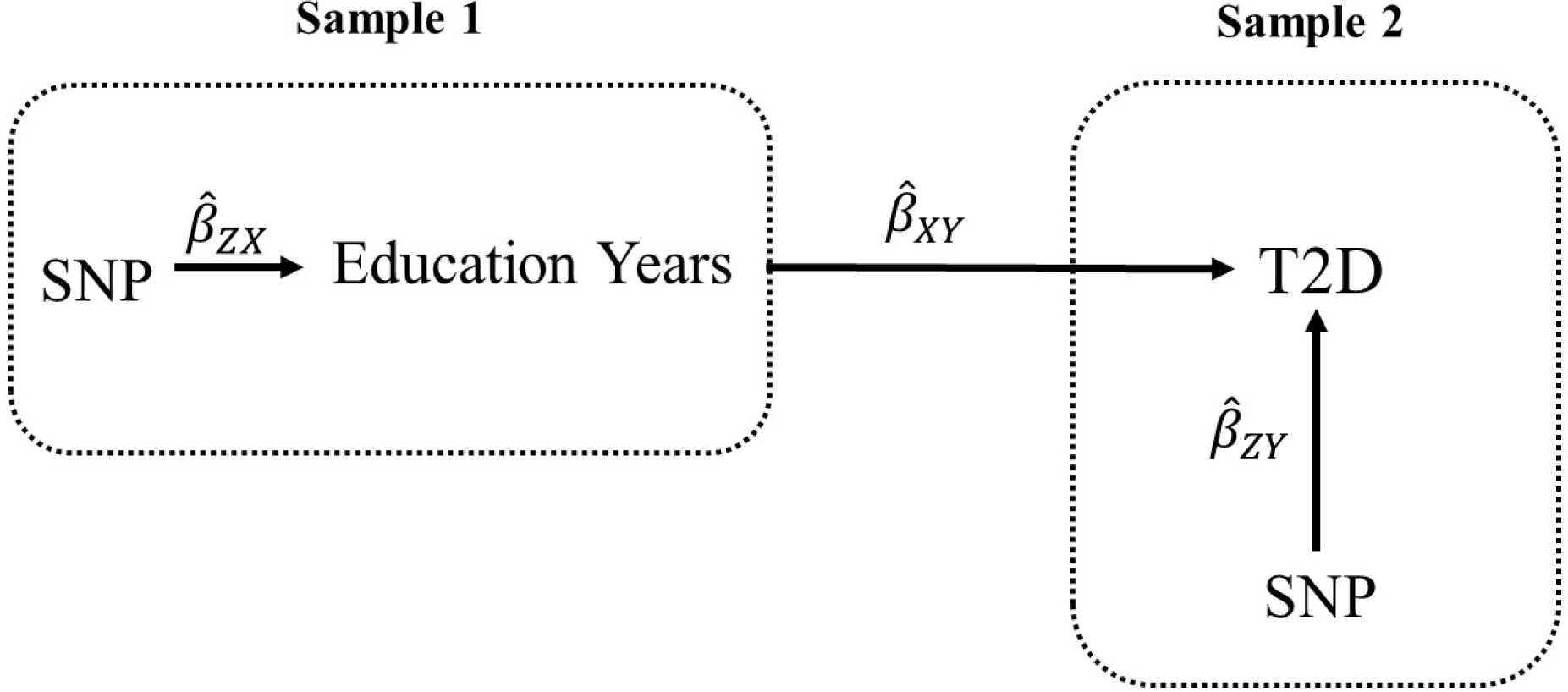
Two-sample MR testing the causal effect of Education Years on T2D. Estimates of the SNP-Education Years associations (*β*^ZX) are calculated in sample 1 (from a genome-wide association, GWA, study of Education Years). The association between these same SNPs and T2D is then estimated in sample 2 (*β*^ZY) (from a T2D GWA study). These estimates are combined into Wald ratios (*β*^XY=*β*^ZY/*β*^ZX). The *β*^XY estimates are meta-analyzed using the inverse-variance weighted analysis (*β*^IVW) method and various sensitivity analyses. The IVW method produces an overall causal estimate Education Years on T2D.

MR also exploits vertical pleiotropy. For example, the assumption that the genetic variants for Education Years have an influence on T2D through their influence on the Education Years is an exploitation of vertical pleiotropy. But vertical pleiotropy is not the only type of pleiotropy. Horizontal pleiotropy also occurs. An example of horizontal pleiotropy would be if the SNPs associated with Education Years were also associated with some other trait (such as socioeconomic status), which then affects risk for T2D. This scenario would constitute a violation to the MR assumptions.

### MR assumptions

MR has the following assumptions: (i) genetic instruments are strongly associated with the exposure; (ii) genetic instruments are independent of confounders of the exposure and the outcome; and (iii) genetic instruments are associated with the outcome only through the exposure^18,20^. For example, the following must be true in order for the present analysis to be valid: (i) genetic variants robustly associated with Education Years must be chosen as instruments to test the causal relationship between Education Years and T2D; (ii) the genetic variants chosen to instrument Education Years must not be associated with confounders of the relationship between Education Years and T2D; and (iii) the genetic variants chosen to instrument Education Years must only impact T2D through their impact on Education Years. When violated, assumption (iii) describes horizontal pleiotropy, which can invalidate causal inference from vertical pleiotropy. Statistically based sensitivity estimators have been developed to evaluate potential violations to assumption (iii) (for more on this, see the subsection, Sensitivity analyses.)

### Design

This study explores the impact of Education Years on T2D and 10 risk factors for T2D. For the later, a list of established risk factors for T2D was obtained from the website for the American Diabetes Association (ADA) (https://www.diabetes.org/diabetes-risk)^21^:

- Being 45 or older
- Being Black, Hispanic/Latino, American Indian, Asian American, or Pacific Islander
- Having a parent with diabetes
- Having a sibling with diabetes
- Being overweight
- Being physically inactive
- Having high blood pressure
- Having low high-density lipoprotein (HDL) cholesterol
- Having high triglycerides
- Having had diabetes during pregnancy (gestational diabetes)
- Having been diagnosed with Polycystic Ovary Syndrome

Of these risk factors, all but “being 45 and older” and “being Black, Hispanic/Latino, American Indian, Asian American, or Pacific Islander” were suitable for investigation with two-sample MR.

### Exposure data source: Education Years

The instrument for Education Years was obtained from a GWA study of Education Years performed by Okbay *et al*. (2016), which included 293,723 participants of European ancestry and adjusted for 10 principal components, age, sex, and study-specific controls^22^. Education Years, inclusive of college, was measured for those who were at least 30 years of age. International Standard Classification of Education (ISCED) categories were used to impute a years-of-education equivalent (SNP coefficients per standard deviation, SD, units of years of schooling; an SD-unit of schooling=3.6 years).

### Outcome data source: T2D

The outcome data for T2D was extracted from Morris *et al*. (2012), which performed a GWA study of T2D in 149,821 participants of European decent, of which 34,840 had T2D^23^. Their GWA adjusted for study-specific covariates and population structure.

### Outcome data source: sibling with diabetes

The outcome data for having a sibling with diabetes was extracted from a GWA study performed by the Medical Research Council-Integrative Epidemiology Unit (MRC-IEU) staff, using PHESANT-derived^24^ UK Biobank data^25,26^ (UK Biobank data field 20111). Briefly, the UK Biobank is an open-access cohort that enrolled about 500,000 participants, largely of European descent^27^. Genetic, health, and demographic data were collected on many of the participants and were made publicly available for researchers. The MRC-IEU staff ran numerous GWA studies with UK Biobank variables, adjusted for age at recruitment and sex, and made their results available through MR-Base, a public repository of summary statistics from GWA studies for use in MR studies. The GWA study of having a sibling with diabetes contained 362,826 participants, of which 31,073 were classified as having a sibling with diabetes.

### Outcome data source: mother with diabetes

The outcome data for having a mother with diabetes was extracted from a GWA study performed by the MRC-IEU staff, which used PHESANT-derived UK Biobank data (UK Biobank data field 20110). The GWA study contained 423,892 participants, of which 40,091 were classified as having a mother with diabetes.

### Outcome data source: father with diabetes

The outcome data for having a father with diabetes comes from a GWA study performed by the MRC-IEU staff, which used PHESANT-derived UK Biobank data (UK Biobank data field 20107). The GWA study contained 400,687 participants, of which 38,850 were classified as having a father with diabetes.

### Outcome data source: overweight status

The outcome data for overweight status come from Berndt *et al*. (2013), which performed a GWA study of clinically defined overweight status in 158,855 participants of European ancestry, of which 93,015 were classified as overweight^28^. Overweight case status was defined as BMI ≥25 kg/m^2^.

### Outcome data source: physical activity

The outcome data for physical activity come from a GWA study by the MRC-IEU staff, which used PHESANT-derived UK Biobank data for moderate physical activity, defined as the number of days of moderate physical activity per week performed for more than 10 minutes at a time. The GWA study included 440,266 participants.

### Outcome data source: high blood pressure

A GWA study of high blood pressure (a binary measure) was performed by the MRC-IEU staff using PHESANT-derived variables^24^ constructed from the UK Biobank data^25,26^ (data field 6150: “Vascular/heart problems diagnosed by doctor: high blood pressure”). There were 461,880 participants, of which 124,227 had high blood pressure as determined by a physician.

### Outcome data source: gestational diabetes

The GWA study of gestational diabetes (a binary measure) was performed by MRC-IEU staff using PHESANT-derived variables ^24^ constructed from UK Biobank data^25,26^ (data field 4041). Participants were asked if they only had diabetes during pregnancy. There were 462,933 participants, 240 of which self-reported having had gestational diabetes.

### Outcome data source: polycystic ovarian syndrome

The outcome data for polycystic ovarian syndrome (a binary measure) was performed by MRC-IEU staff using PHESANT-derived variables^24^ constructed from the UK Biobank data^25,26^ (data field 20002). There were 462,933 participants, of which 571 self-reported having polycystic ovarian syndrome.

### Outcome data source: HDL levels

The outcome data for circulating HDL levels (a continuous measure) come from Willer *et al*. (2013), which performed an age- and sex-adjusted GWA study of circulating HDL levels in up to 187,167 individuals, largely of European ancestry^29^.

### Outcome data source: triglyceride levels

The outcome data for triglyceride levels (a continuous measure) come from Willer *et al*. (2013), which performed an age- and sex-GWA study of circulating triglyceride levels in up to 177,861 individuals, largely of European ancestry^29^.

To ease interpretability, all MR results for the effects of Education Years on T2D and T2D risk factors were exponentiated from log odds to odds ratios, except for outcomes of continuous variables (i.e., HDL and triglyceride levels), which are presented as beta estimates (Table 1).

**Table 1.**
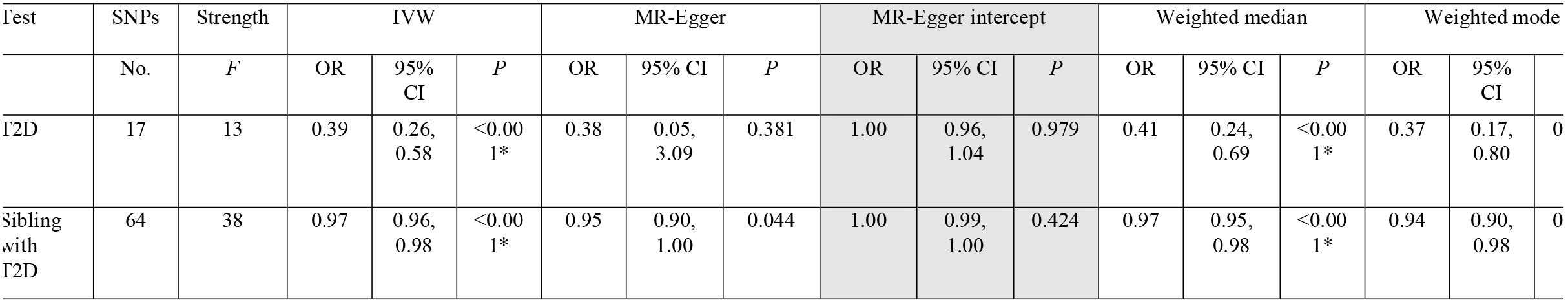
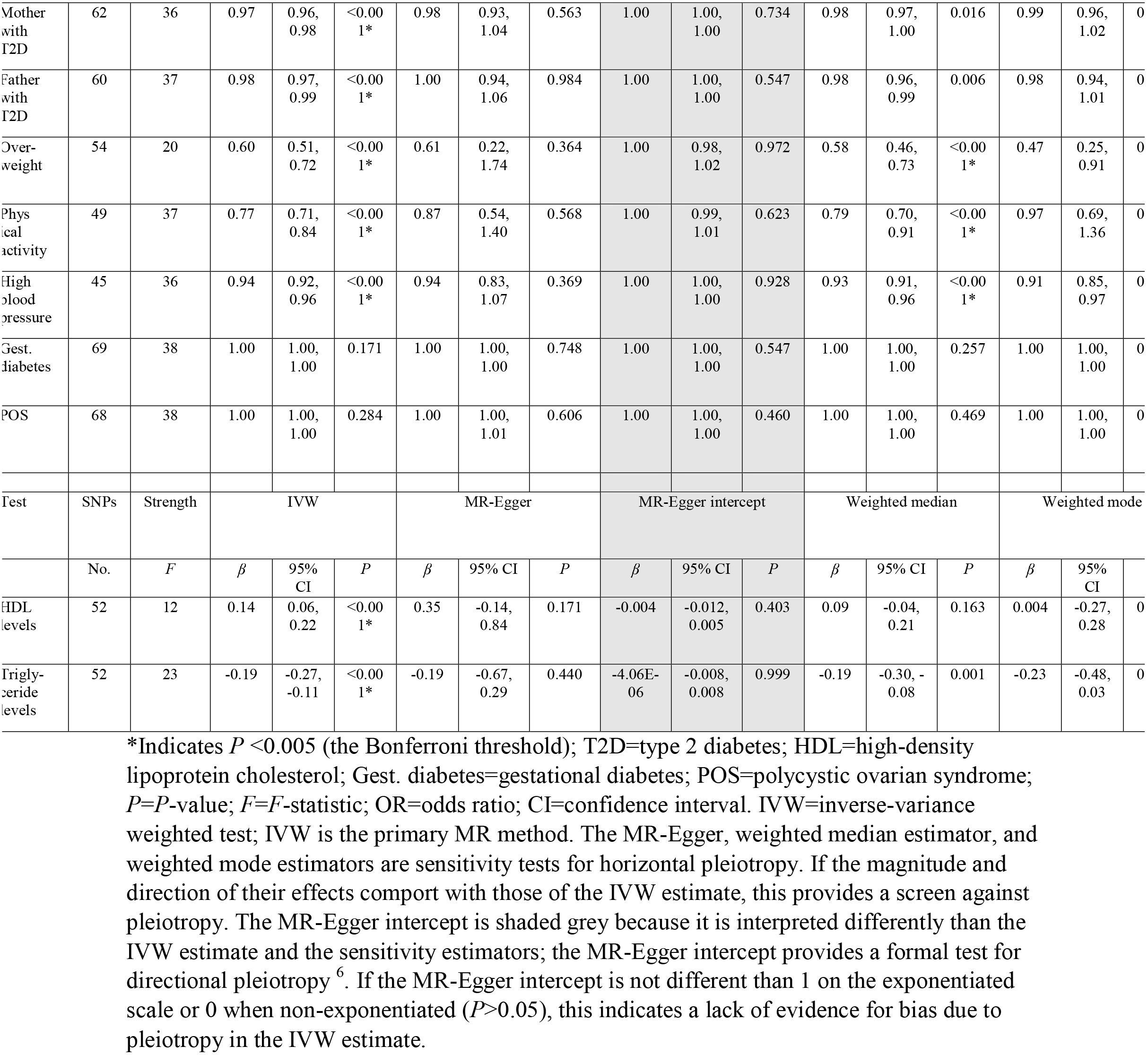
Causal estimates for Education Years on T2D and 10 risk factors for T2D.

The summary statistics used for the MR analyses are available in Supplementary Tables 1-11.

### Instrument construction

As introduced in Figure 1, independent (those not in linkage disequilibrium, LD; R^2^ < 0.01) SNPs associated at genome-wide significance (*P* < 5 × 10^−8^) with Education Years were extracted from the Okbay *et al*. (2016) GWA study. The summary statistics for the Education Years-associated SNPs were then extracted from each of the outcome GWA studies. SNP-Education Years and SNP-outcome associations were harmonized and combined with the IVW method (**Figure 1**).

### Sensitivity analyses

A weakness of the IVW estimator is that its estimate can be biased if the meta-analyzed SNPs are directionally pleiotropic^30^. This can cause a violation to MR assumption (iii) and invalidate the findings. To address this, MR-Egger regression, weighted median, and weighted mode MR methods can be run as complements to the IVW. The directions and magnitudes of their effect estimates can be compared to those of the IVW. Doing so is a type triangulation: comparing approaches that have different assumptions to weigh evidence^31^. The reason for this is that the various MR sensitivity estimators make different assumptions about possible underlying pleiotropy. Due to their different assumptions, it is unlikely that the IVW and sensitivity estimators would be homogeneous in the directions and magnitudes of their effect estimates if there were substantial violations to MR assumption (iii). Therefore, triangulating their directions and magnitudes of effects provides a screen against pleiotropy. (Nuanced descriptions of how the various MR estimators deal with pleiotropy are described elsewhere^30,32,33^). MR-Egger regression, weighted median, and weighted mode MR sensitivity methods were run for all analyses.

A formal test for directional pleiotropy was also done with the MR-Egger intecept. If the MR-Egger intercept is not different than 1 on the exponentiated scale or 0 when non-exponentiated (*P*>0.05), this indicates a lack of evidence for bias due to pleiotropy in the IVW estimate.

In addition, potential outlier SNPs were removed using RadialMR regression^34^ for the MR tests of Education Years on T2D risk factors. (The differing number of SNPs for the Education Years instruments is due to this and that the various outcome GWA studies not having a uniform set SNPs in their association studies). All instrumental variables included in this analysis have Cochrane’s *Q*-statistic *P*-values indicating no evidence for heterogeneity between SNPs^35^. Heterogeneity in the effect estimates for SNPs can indicate pleiotropy. Thus, ensuring a lack of heterogeneity between SNPs is an additional method to boost the chance that MR assumption (iii) is not violated. Heterogeneity statistics are provided in Supplementary Tables 12-22.

The IVW and sensitivity estimations were performed in R version 3.5.2 with the “TwoSampleMR” package^16,36^. Overall, 11 tests were performed. The Bonferroni correction was used to penalize for multiple testing: *P*=0.05/11 (0.005).

### Power

The study was powered for the test of Education Years on T2D, using mRnd MR power calculator (available at http://cnsgenomics.com/shiny/mRnd/)^37^. There was ≥80% power to detect odds ratios in the range of 0.3-0.7 (Figure 2). In addition to the overall power to detect an association, MR studies also rely on *F*-statistics. *F*-statistics provide an indication of instrument strength^38^. *F*-statistics <10 are conventionally considered to be weak^39^. *F*-statistics for each test are available in Table 1.

**Figure 2.**
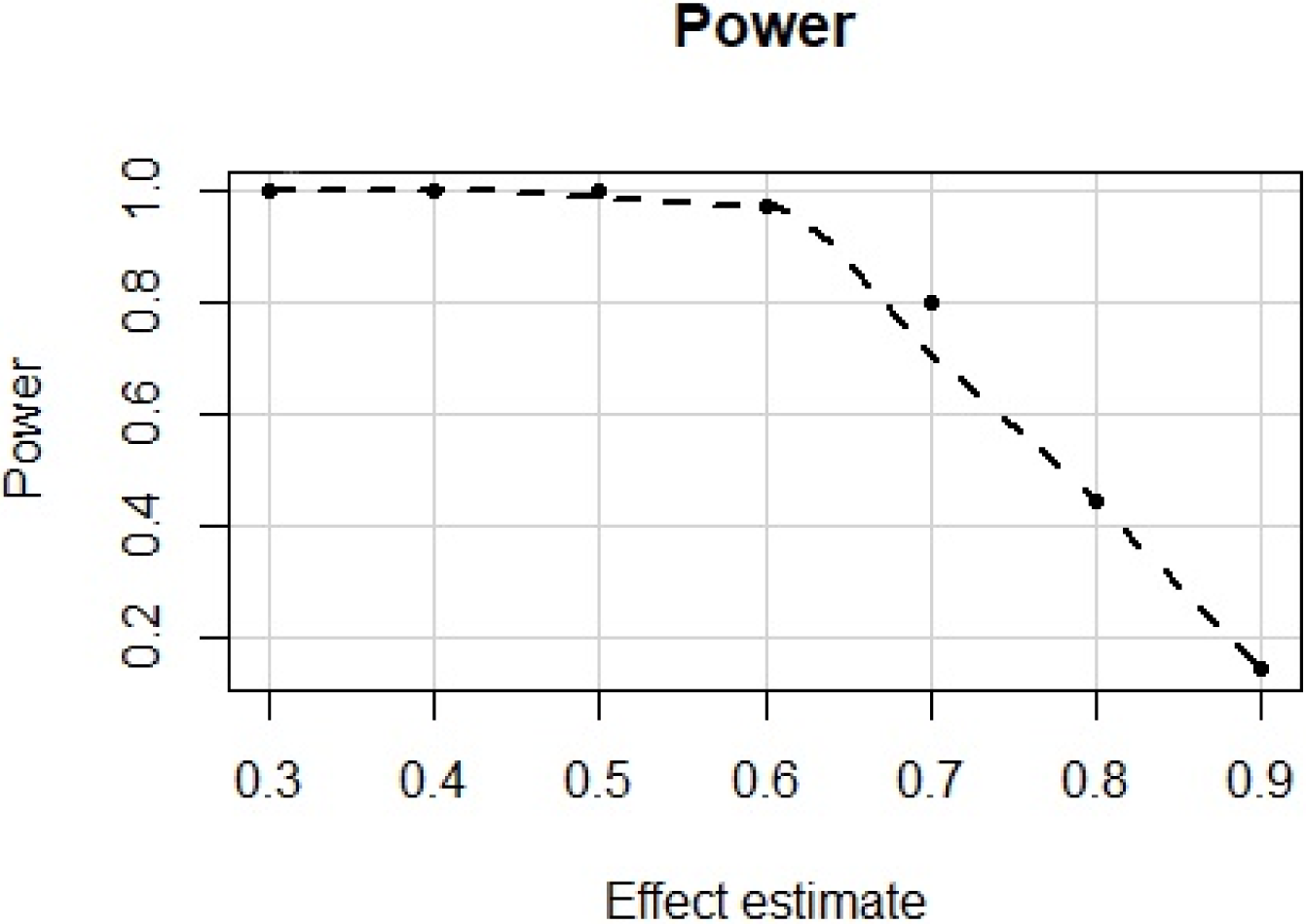
Power calculations for a range of plausible effects estimates for the MR test of Education Years on T2D.

## Data Availability

Data availability. All data sources are publicly available and are accessible within MR-Base: http://www.mrbase.org/.

## Data availability

All data sources are publicly available and are accessible within MR-Base: http://www.mrbase.org/^16^.

